# Predicting human health from biofluid-based metabolomics using machine learning

**DOI:** 10.1101/2020.01.29.20019471

**Authors:** Ethan D. Evans, Claire Duvallet, Nathaniel D. Chu, Michael K. Oberst, Michael A. Murphy, Isaac Rockafellow, David Sontag, Eric J. Alm

## Abstract

Biofluid-based metabolomics enables the profiling of thousands of molecules and has the potential to provide highly accurate, minimally invasive diagnostics for a range of health conditions. However, typical metabolomics studies focus on only a few statistically significant features. We study the applicability of machine learning for health state-prediction across 35 human mass spectrometry-based metabolomics studies. Models trained on all features outperform those using only significant features and frequently provide high predictive performance across nine health states, despite disparate experimental conditions and disease contexts. Combining data from different experimental settings (e.g. sample type, instrument, chromatography) within a study minimally alters predictive performance, suggesting information overlap between different methods. Using only non-significant features, we still often obtain high predictive performance. To facilitate further advances, we provide all data online. This work highlights the applicability of biofluid-based metabolomics with data-driven analysis for health state diagnostics.

---

While fundamental to personalized healthcare, it is often challenging to diagnose an individual’s health state—a general term encompassing disease and non-disease phenotypes like age—due to low test sensitivity, specificity or the requirement of invasive procedures. Body-fluid sampling (e.g. blood or urine) offers a minimally invasive approach to identify health conditions throughout the body. The traditional concept of biofluids-based diagnostics relies on health-state biomarkers. Biomarkers cover a broad spectrum of measurements^1^, but typically refers to a small number of select and specific molecules or biopolymers, capable of differentiating healthy from diseased states. Currently, many biomarker-containing tests are used in routine lab monitoring (e.g. complete blood count, ‘basic’ and ‘comprehensive’ metabolic panels, lipid panels, etc.) providing coarse health-state categorization. Disease-specific tests exist, displaying a range of sensitivity and specificity—including apolipoprotein E, with other measurements, for Alzheimer’s diagnostics^2^, the prostate-specific antigen test for prostate cancer^3^, alpha fetoprotein (AFP) for liver cancer^4^, as well as a recent use of the SOMAscan^5^ for diagnosing tuberculosis^6^.

Metabolomics rapidly supplies information on thousands of molecules, and represents a method for biofluid-based diagnostics^7–9^. To date, serum, plasma, urine and cerebrospinal fluid (CSF) metabolomics has been applied to many health states, ranging from cancers^10–13^ and infectious diseases^14,15^ to Chronic Obstructive Pulmonary Disease (COPD)^16^, smoking^17^ and Alzheimer’s disease^18,19^. These studies are regularly performed using analytical instrumentation like liquid or gas chromatography mass spectrometry (LC-MS and GC-MS respectively) as well as nuclear magnetic resonance (NMR)^20^. Frequently, the goal of such studies is to determine the chemical identity of the features that are significantly more or less abundant between health states, where a ‘feature’ is a molecule defined by a mass-to-charge ratio (mz) and retention time (rt). While the majority of features often remain unidentified, analysis of the chemically identified features allows for biological or translational interpretation. A common approach to understanding the biological perturbations underlying disease is to monitor changes in metabolite levels and map compounds to biochemical pathways^18,21^. Many studies then leverage select chemically identified features to perform differential diagnostics, predictive modeling, or to analyze health state association. For instance, one study found select amino acid metabolism associated with later development of diabetes^22^; another studied populations in China, Japan, the UK, and the USA and uncovered metabolites associated with blood pressure^23^.

Mass spectrometry (MS) based diagnostic models typically do not use the full set of features, reducing the high-dimensional MS data to a limited set of hypothesis-generating, usually identified, features^17,19^. For binary classification, univariate statistical tests (Student’s t-test or Mann-Whitney U-test, MW-U) are routinely used to isolate statistically significant features—usually identified using false discovery rate (FDR) adjusted P-values < 0.05. These significant features are then used by algorithms like partial least squares discriminant analysis (PLS-DA) for both additional dimensionality reduction and health-state classification. Additional classification methods employed include random forests^24–26^ and support vector machines^27^. Other analyses are strictly based on univariate or multivariate receiver operating characteristic-area under the curve (ROC-AUC)^26,28^. In addition to standard statistical methods, alternative means of feature selection are often performed: feature enrichment^29^, manual and statistical curation^30^, and the use of discovery cohorts to suggest metabolites for targeted analysis and model development^19,31^.

We aimed to determine whether complete metabolomics data sets, combined with machine learning (ML) models, could provide robust diagnostic performance across nine general health state categories. We performed standardized data processing and analysis on a set of 35 publicly available, predominantly untargeted LC- or GC-MS studies. In total, we analyzed 148 individual MS data sets from diverse experimental conditions, including sample type, chromatography method, and MS ionization mode. Diagnostic classification was performed using an interpretable ML model: logistic regression with L1 regularization (L1-LR). This regularization technique induces sparsity on the set of features used by the model to perform diagnosis, reducing dimensionality. As we were primarily interested in diagnostic performance, we bypassed the challenging problem of chemical identification and focused solely on feature characteristics (i.e. the intensity values of specific mz and rt pairs). We found that models trained on all features demonstrated high predictive power and outperformed those that used only statistically significant features. Moreover, models often achieved high classification performance using only non-significant features, demonstrating the predictive information contained in a complete data set. These results were observed across all biofluid types and analytical methods, with the major determinant of performance being which disease or health state analyzed. Of the nine health-state categories, cancer was the most challenging to diagnose and displayed the largest range of model predictive performance. In contrast, health states including infectious diseases, cardiovascular, and rheumatologic states, showed high model predictive performance. These results suggest that biofluid-based metabolomics are a promising technology for health-state diagnostics.

## Results

### Biofluid-based metabolomics studies cover many health states with various cohort sizes

Literature and database searches identified 35 studies, covering nine general health state categories (Figure 1A). While one lung cancer study^25^ provided data for 1005 individuals, most studies included tens to a few hundred individuals. Cancer represented the largest category, with 14 studies^11,12,25,29,31–39^; cardiovascular^28^, rheumatologic^26,‡^, renal^‡^, and respiratory^16^ diseases were represented by only one or two studies. Other health states with a small to medium number of studies included neuro/neuropsychiatric^18,19^, endocrine^21,24,40,41^, and infectious disease^14,15,27,30,42,43^, as well as a general ‘other’ category^17,44–46^.

**Figure 1.**
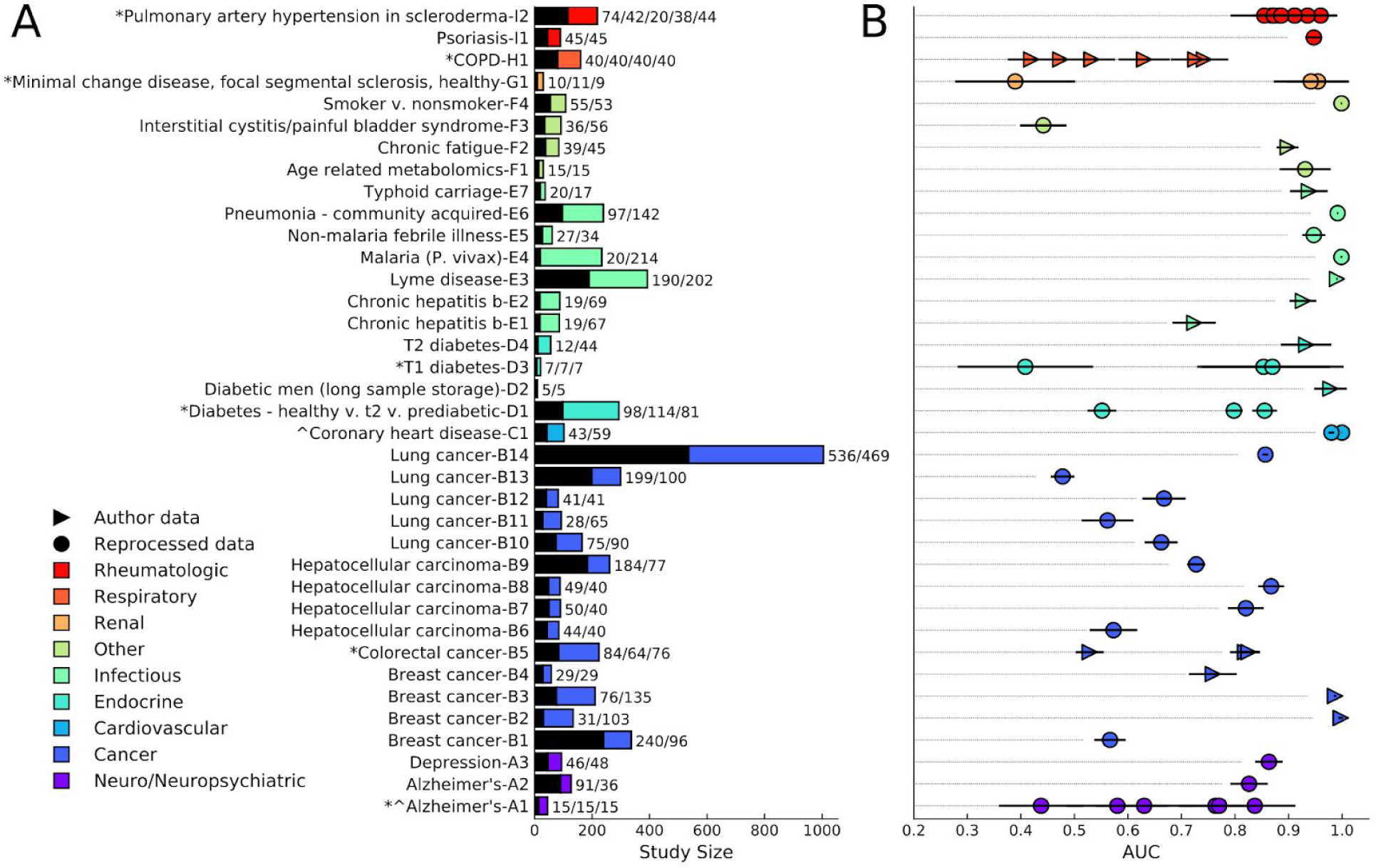
Body fluid based metabolomics often possesses health state-dependent signal and diagnostic capability. (A) Studies analyzed and their associated cohort sizes (control/case sizes to the right of the bar plot), separated by health state category. Shown in black are the controls, with the cases in color. Multiclass control bars correspond to the size of the first class for 3-class studies, or first 2 classes of 4- or 5-class studies, with the case bar representing the remaining samples. Multiclass studies are: D1 (Type 2 diabetes, prediabetic, healthy), H1 (never smokers, former smokers, smokers, COPD patients), D3 (Type 1 diabetes—insulin injection, Type 1 diabetes—insulin withdraw, no diabetes), A1 (Alzheimer’s, mild cognitive impairment, normal), B5 (colorectal cancer patients, polyp patients, healthy controls), G1 (minimal change disease, focal segmental glomerulosclerosis, control), I2 (normal, pulmonary artery hypertension, low risk, healthy, borderline pressure). (B) Averaged ROC-AUC and standard deviation analysis for 30 L1-LR models, each trained and tested on different randomized, stratified shuffles of the within-study combined data sets. *Multiclass studies for which one-vs-one models were built. ^Studies for which it was not possible to combine data sets.

We reduced complex studies to single or multiple binary classification problems, typically between a control state and disease or altered health state. As an example, Alzheimer’s study A2^19^ had a complex discovery cohort and targeted validation set that looked at differences between healthy controls, individuals before and after conversion to a cognitively impaired state, and individuals who entered the study with cognitive impairment. For this study, we analyzed the untargeted mass spectrometry (MS) data, which included only healthy and pre-study cognitively impaired individuals.For one breast cancer study^38^, we analyzed the serum (B2) and plasma (B3) data separately, as the study included differently sized discovery and validation cohorts with different sample types. The seven multiclass studies were analyzed via multiple one-to-one comparisons as done in many metabolomics studies (Figure 1A). The chronic hepatitis B study^15^ was split in two because the targeted oxylipin assay (E1) possessed a different number of cases from the untargeted lipid analysis (E2). One breast cancer study^39^ (B1, Table S1) aimed to generate prognostic predictions for response to chemotherapy among patients with tumors.

### Biofluid profiles enable health state prediction using a data-driven approach

Metabolomics features from LC- and GC-MS studies across the nine health states generally provided moderate to high predictive performance (Figure 1B). When raw MS data was available, we reprocessed it using an in-house pipeline, generating feature tables of peak intensities associated with each mz-rt pair (referred to as ‘reprocessed’ data sets, *Methods*). If only preprocessed feature tables were available, all features were used (referred to as ‘author’ data sets); however, these were not always complete, unfiltered data sets. To minimally bias our conclusions and treat all data in a similar manner, each data set was percentile normalized^47^. For each study with multiple data sets, we combined all data into a single data set (e.g. concatenating the positive and negative ionization mode data from an LC-MS study) when possible; for two studies^18,28^ it was not possible to match samples across data sets. To establish baseline predictive performance, we trained L1-regularized logistic regression (L1-LR) models for each study (*Methods*). Many displayed test AUC values > 0.7 for at least one comparison (Figure 1B). For most multiclass studies, depending on which one-versus-one health state comparison was performed, we observed near random guessing (AUC ∼ 0.5) or high AUC values (> 0.7). Nearly all of the infectious disease models displayed significant predictive power, even for studies with small cohorts. Rheumatologic, cardiovascular, neuro/neuropsychiatric, and select ‘other’ health states, similarly, possessed high AUC values. In contrast, seven out of 14 cancer studies displayed low model performance (< 0.7), and the interstitial cystitis/painful bladder syndrome study yielded random model guessing.

### Individual data sets from different experimental conditions display mixed predictive performance

Models trained on individual data sets within a study generally performed well across different experimental conditions, including biofluid type, method of chromatographic separation, MS instrumentation, and ionization mode. Plasma and serum accounted for the vast majority of studies, with only four using urine, two using CSF, and one using dried blood spots (DBS, Figure 2A). We found that sample type did not significantly affect model performance, with AUC values predominantly reflecting the disease or health state under study. Likewise, plasma- and serum-based studies displayed no statistically significant difference in test set AUCs (Figure 2B and Figure S1). For the chronic hepatitis B study, models built using the untargeted metabolomics data (E2), in both positive and negative ionization mode, substantially outperformed the targeted oxylipin assay (E1, Figure 2B). When analyzing studies with only two classes, LC outperformed GC (P = 0.0014 MW-U test, Figure 2C). However, a disproportionate number of GC data sets (12 of 19 as opposed to 10 of 37 for LC) were from cancer studies, the most challenging diagnostic category. Further, LC-based studies generally possessed 1–2 orders of magnitude more features for model training (Figure S2). Most studies used either only positive or both MS ionization modes; only two used solely negative mode, both of which displayed test AUCs > 0.7 for at least one comparison. For binary class LC-MS data sets, ionization mode and column type did not appear to alter predictive performance (Figure 2C). These results may be biased by relatively small sample sizes in addition to study- or health state-specific experimental parameters.

**Figure 2.**
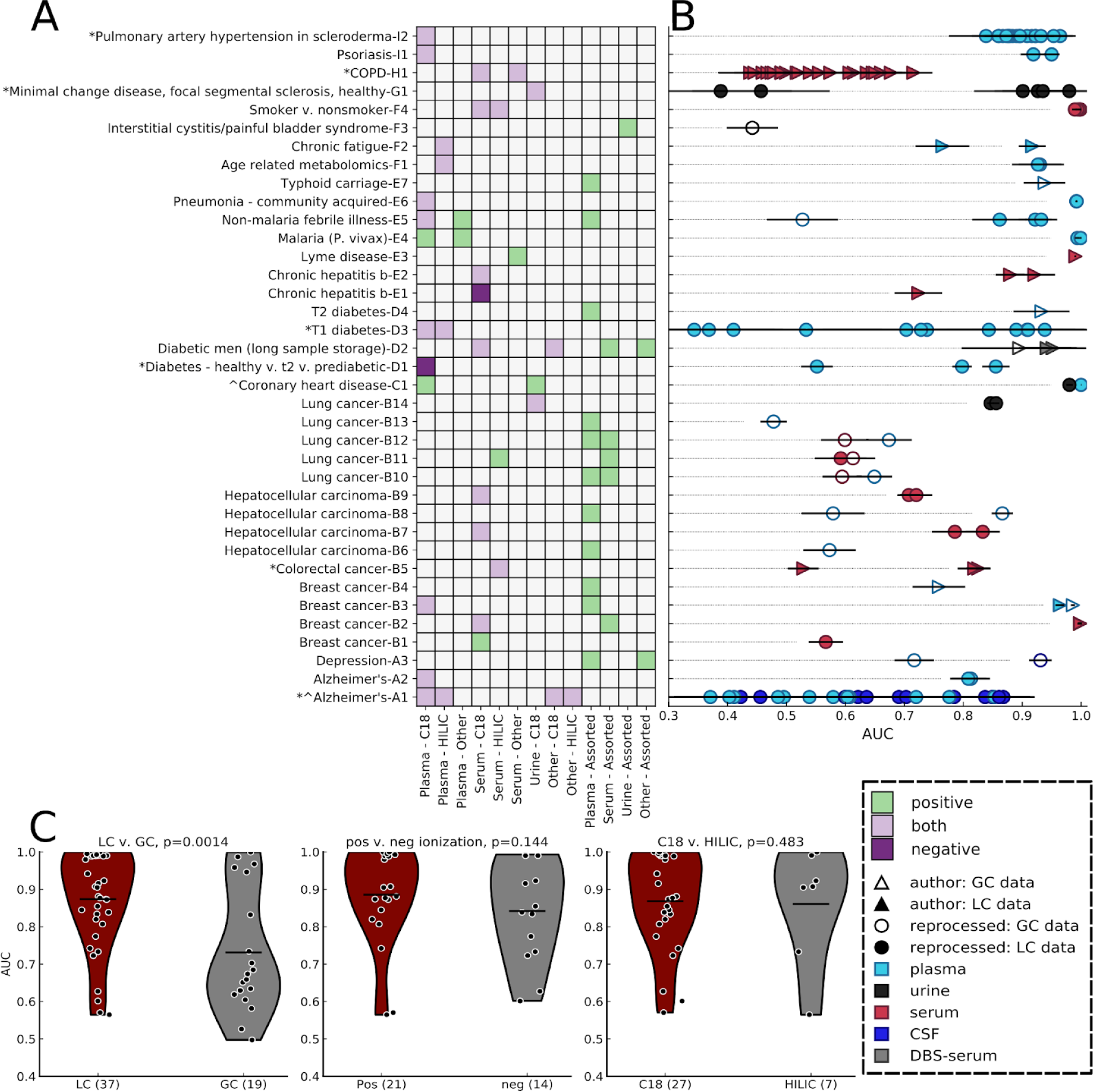
Health state information is found in all body fluids using different instruments, mass spectrometry ion modes, and chromatographic methods. (A) Ion mode, sample, and column type (for liquid chromatography) used for each individual data set. (B) Individual data set ROC-AUC and standard deviation analysis of 30 averaged L1-LR models, each trained and tested on different randomized, stratified shuffles, with associated sample and instrument type.(C) Violin plots for the comparison of non-multiclass, liquid chromatography to gas chromatography data sets (left), positive versus negative ion mode for all liquid chromatography data sets (middle), and C18 versus HILIC columns (right), using AUC values from models trained on individual data sets, using all features.

### Biological considerations may explain select low performance models

Several low performing models originated from multiclass studies—specifically, comparisons between similarly presenting health states. Low AUC values were seen in studies on Type 1 diabetes, Type 2 diabetes, Alzheimer’s, colorectal cancer, COPD, and one study on two nephrotic syndromes: minimal change disease (MCD, a kidney disease characterized by significant urine protein levels) versus focal segmental glomerulosclerosis (FSGS, scarring of the kidney that may also present with high protein levels in urine, Figure S3). For instance, in the colorectal cancer study, it was not possible to distinguish between healthy individuals with non-cancerous polyps versus those without (AUC = 0.529 ± 0.009, mean and 95% confidence interval respectively). However, we were able to differentiate true cancer cases from both healthy states (AUCs = 0.819 ± 0.010, 0.825 ± 0.007). Similarly, differentiating between MCD and FSGS was not possible (AUCs = 0.388 ± 0.136 and 0.457 ± 0.132 for positive and negative ionization LC-MS, respectively), yet both were easily distinguished from healthy controls (AUCs > 0.9). This pattern extended to other health states with multi-class studies (Figure S3). These cases highlight the difficulty of differentiating health states with similar metabolic signals, something that appears increasingly common as the number of classes increases.

### Using all features, not solely statistically significant features, provides the best performance

We next assessed the performance of L1-LR models using two separate data set modifications. The first tested whether there is more predictive power when using data combined from different experimental conditions (as done for Figure 1B). We observed that the combined data set models generally performed on par with the average of models built using individual data sets (average AUC increase of 0.026, not statistically significant P = 0.219), with few combined data set models showing increased performance (Figure 3A). This result illustrates that larger feature sets, putatively with more molecular information, may not lead to improved diagnostic performance. The minimal increase in performance further suggests that data from different experimental conditions contains redundant information. For this reason, we chose to focus our analysis on models trained on individual data sets.

**Figure 3.**
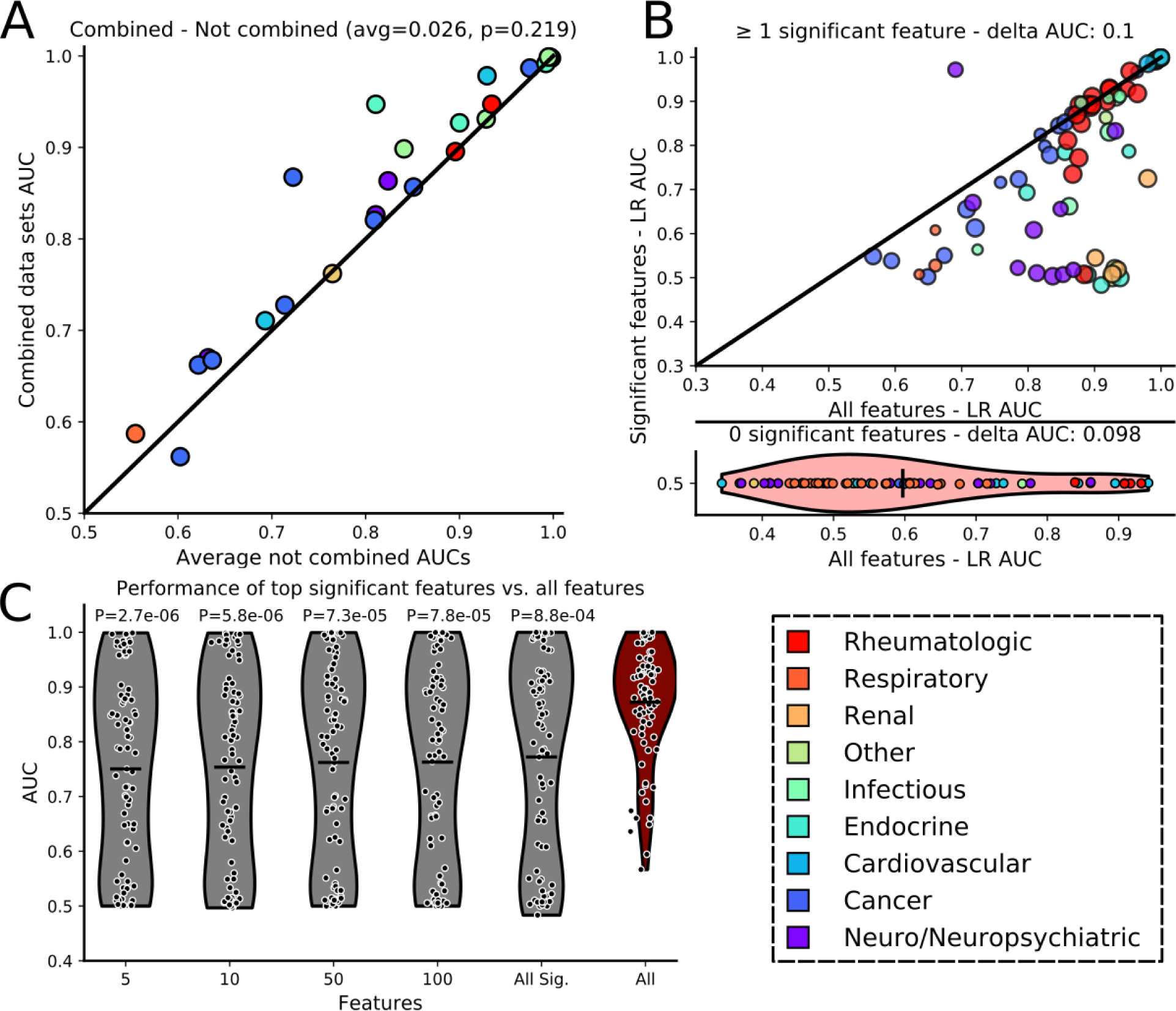
Diagnostic performance is primarily improved by using all features, rather than using only statistically significantly features or combined data sets. (A) Comparison of AUC values from L1-LR models built from within-study combined data sets versus the average AUC of independent models built on non-combined data sets.The average difference between combined and non-combined model AUCs, along with the P-value for the comparison (MW-U test). (B, top) Comparison of model performance on individual data sets with at least one statistically significant feature. Circle size is proportional to the number of features. The purple outlier point with better AUC using only significant features is from a HILIC, CSF sample in positive ionization from study A1. (B, bottom with shared y-axis label) Comparison of performance between models trained using all features versus ‘significant feature only’ models when there were no significant features with accompanying violin plot of the AUC distribution of models built using all features. Delta AUC values were determined by subtracting the AUC of a model built with only significant features from that of a model trained on all features, and then averaged over all data sets. (C) Comparison of models built using up to 5, 10, 50 and 100 of the most significant features (lowest Q-value) or all significant features, versus models trained using all features; results displayed only for data sets that possessed significant features. P-values correspond to MW-U tests between the AUC values from models using all features versus the AUC values from models built with a select number of significant features. Health state color legend for (A) and (B) shown in the bottom right.

The second modification tested the effect of reducing the feature space to only statistically significant features (FDR corrected P-values < 0.05). We split data sets into two groups: those with at least one significant feature during training, and those without. The latter, lacking features to train on, were given an AUC of 0.5, representing random guessing (Figure 3B and *Methods*). For data sets with significant features, 75% of models trained using all features outperformed those that used only statistically significant features (Figure 3B, top). For data sets lacking significant features, models trained with all features, unless overtrained, generally outperformed the “random guessing” models (assigned AUC = 0.5), showing an approximately 0.1 average increase in AUC (Figure 3B, bottom).

Observing improved performance in models trained using all features, we tested whether the class of model affected performance. To ground our study with a commonly used model, we compared the performance of L1-LR models to partial least square discriminant analysis (PLS-DA) models, with both using all the available features. The two techniques provided similar performance. However, the L1-LR models may offer increased interpretability as they provide sparse sets of explanatory features (Figure S4).

While using all features appeared optimal, significant features alone did show predictive power. Thus to study their importance and information content we tested model performance on subsets of the most significant features. Using up to 5, 10, 50 or 100 (or all in cases where the number of features was less than one of these values) of the most significant features demonstrated that—for each comparison—the models built with all features outperformed the significant feature-based models (Figure 3C). This analysis excluded the 0.5 AUC-assigned data sets that lacked significant features. However, when these data sets were retained, the difference in model performance remained (Figure S5). For most data sets, using any number of significant features resulted in similar predictive power (Figure S6); demonstrating that a small number of significant features accounted for the majority of predictive power of the significant features.

### L1-LR provides sparse models that use both statistically significant and non-significant features

We examined the extent to which L1-LR models trained on all features across different health states recovered significant features, as well as the degree to which the features that were used (i.e. had non-zero coefficients) were non-significant. We found that the models used a wide range of statistically significant features, from zero to almost all (Figure 4A). While this affirmed the importance of significant features, for many models, non-significant features constituted a large fraction of the features used. In fact, among models trained on data sets with at least one significant feature, AUC only showed partial correlation (R = 0.52) with the fraction of significant features used (Figure S7).

**Figure 4.**
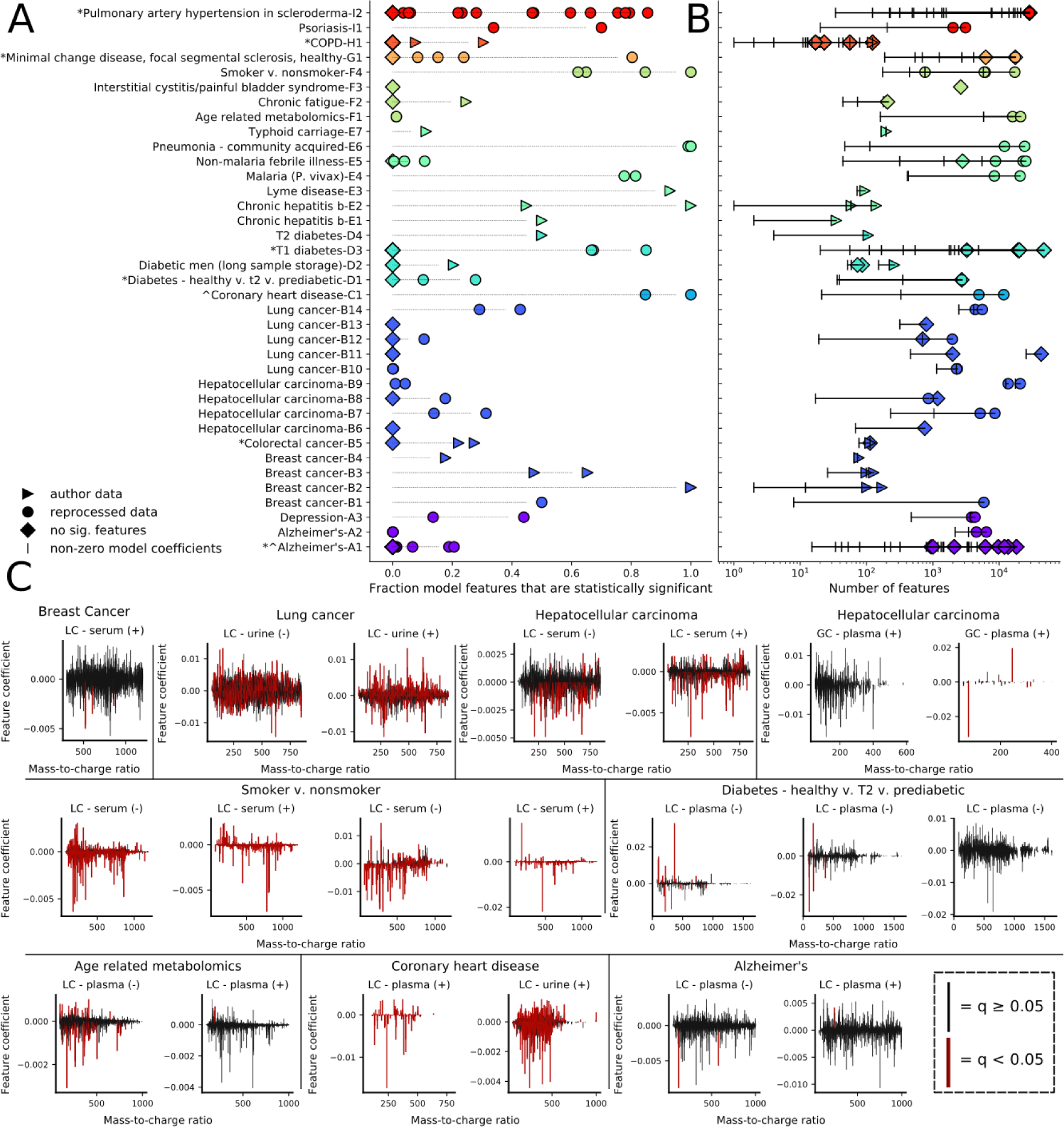
Machine learning models are relatively sparse and frequently use features spanning a large mass range and both significance types. (A) Fraction of non-zero feature coefficients in the models corresponding to statistically significant features (p < 0.05 FDR-corrected, MW-U test) for single models trained on individual data sets. (B) Number of input features (colored points) relative to the number of non-zero model features (vertical dashes) for a single, representative model training for each data set. (C) Representative plots of the features and associated average model coefficients (30 model trainings) used across the range of observed mass-to-charge ratios. Significant versus non-significant features are depicted in different colors (red, q < 0.05; black, q ≥ 0.05). Data sets (clockwise, starting from the upper left): Breast cancer (B1), Lung cancer (B14), Hepatocellular carcinoma (B7), Diabetes (D1), Alzheimer’s (A2), Coronary heart disease (C1), Age (F1), Smoking versus non-smoking (F4).

Relative to the number of input features, L1-LR models trained using all features provided sparse solutions, supplying increased interpretability by implicit feature selection (Figure 4B). Analyzing a single model for each data set, we found that a large range of total features were used (from tens to tens of thousands); but that the number used was generally reduced by an order of magnitude relative to the number of input features (Figure 4B, vertical dashes). This reduced feature space may identify features, notably those with high model coefficients, that are especially important for a given predictive task and follow up analysis. Chemical identification of these features would allow for biochemical and systems-level analysis to better understand these diseases or health states.

### Features with a diverse set of properties are used

Trained models used a relatively large number of features with small coefficients (< 0.005), spanning range of mz and rt values (Figure 4C and Figures S8-S12). Only a handful of features possessed relatively large model coefficients (> 0.005). Select models primarily used significant features (F4), while others used mostly non-significant features or a mixture of both (B1); importantly, in both cases features coefficients could be large or small for either significance type (Figure 4C). Greater model coefficients were often observed for features with larger enrichment factors between cases and controls; however, a majority of features possessed near zero feature coefficients or minimal enrichment, limiting analysis (Figures S13–S21). Across studies, the majority of the mz domain possessed features that were used by the models, putatively illustrating that many different molecules, constituting a substantial portion of the biofluid sample, provided health state information.

### Non-significant features alone can provide high model performance

In light of the improvement observed using all features, we wished to understand the predictive capabilities of only the non-significant features. We removed all significant features from each data set and trained models on the remaining non-significant features. A surprising number (∼51%) of models still achieved high AUC values of ≥ 0.7 (Figure 5A, dashed line). As a reference, 61% of models trained using all features achieved an AUC ≥ 0.7, with an average AUC difference of 0.111 between the two cases. Additionally, we observed high AUC values across multiple health-state categories and experimental parameters (Extended Supplementary Data 1).

**Figure 5.**
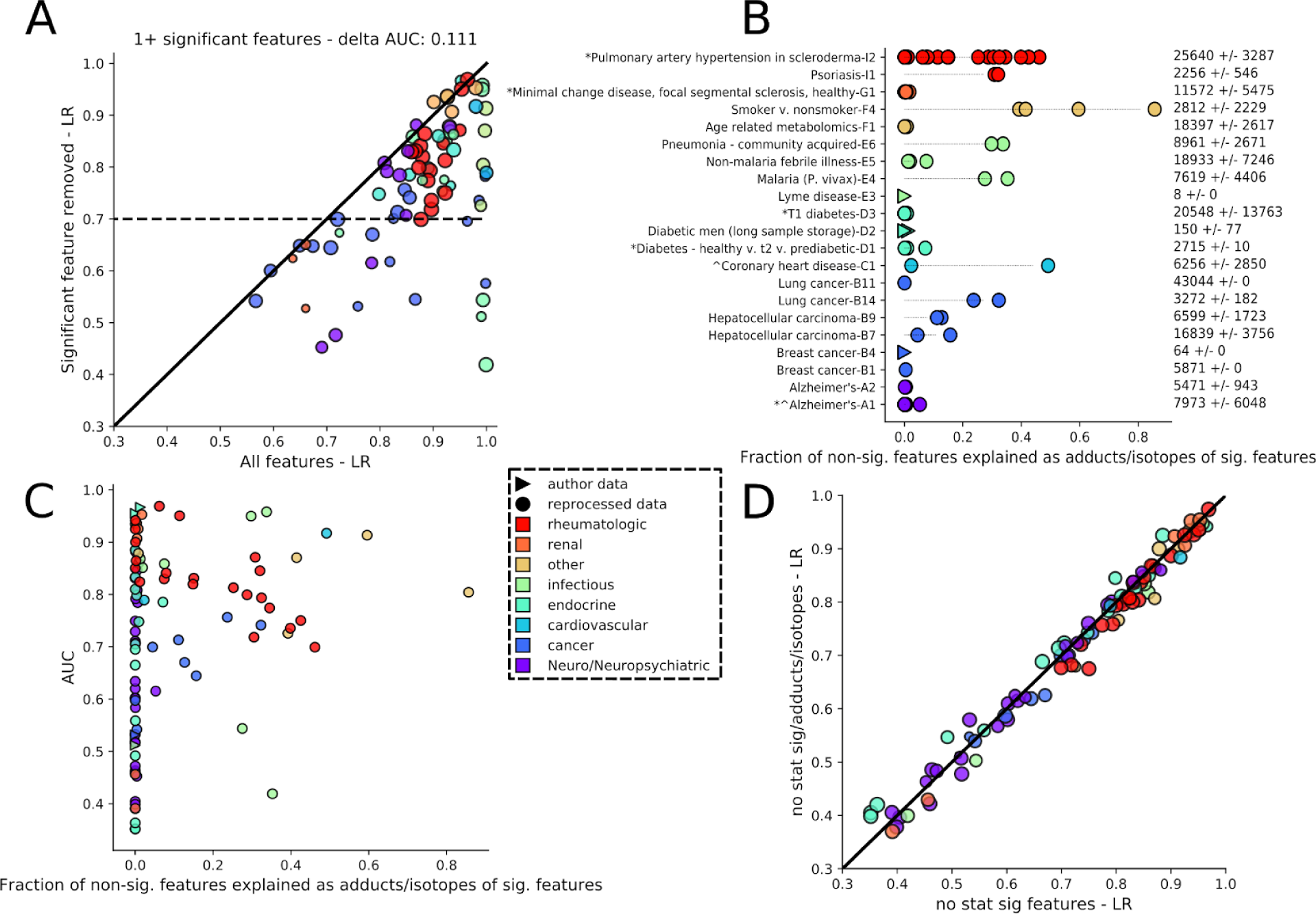
Models trained with only non-significant features often retain relatively high diagnostic performance. (A) AUC comparison between models trained using all features and those using no statistically significant features for data sets with at least one significant feature. Circle size is proportional to the log of the number of features in the data set. (B) Fraction of non-significant features for each high resolution mass spectrometry data set that can be explained as adducts or isotopes of statistically significant features. Numbers on the right are the average number of features across a study’s data sets, with standard deviation. (C) AUC of models trained and tested using only non-significant features versus the fraction of non-significant features explained by adducts and isotopes in the input data set. (D) AUC comparison between models trained using only non-significant features versus non-significant features without adducts or isotopes of significant features. Circle size is proportional to the log of the number of features in data sets from which significant features, their adducts, and isotopes have been removed.

We verified that the performance was not due to information from the significant features remaining in their non-significant isotopes and adducts. For this, we analyzed the high resolution (HR) MS data sets in which isotopes and adducts could be determined. In 9 of the 21 HR-MS data sets, more than 10% of the non-significant features could be explained as putative adducts or isotopes of the significant features (Figure 5B, *Methods*). Furthermore, all 9 displayed AUC values > 0.7. Among data sets with a large number of total features, some had high representation of isotopes and adducts while others had relatively few (Figure 5B). However, low numbers of isotopes and adducts likely arose due to data sets possessing only a small number of significant features (Figure S22). Additionally, data sets with a larger number of significant features tended to come from studies with larger cohorts as opposed to simply having the most initial features (circle sizes, Figure S22). After removing significant feature isotopes and adducts, newly trained models showed a linear relationship with those that included isotopes and adducts (Figure 5D). The features used by these models spanned a similar range of MS intensity values to those used in previous models and were not biased to background (low intensity) features (Figure S23). A similarly wide mz-range of features possessed useful predictive information, despite being non-significant (Figures S24–S29). Like full data set-trained models, a handful of features possessed large absolute coefficient values (0.01–0.4) while many had very small coefficient features (< 0.0005). These results indicate that high AUC values were not simply a result of remnant information from the significant features.

## Discussion

This analysis set out to evaluate the predictive power of biofluid metabolomics for machine learning based diagnostics. In many cases, biofluids provide robust diagnostic capabilities. Here we discuss: (1) health states suited for metabolomics-based diagnostics, (2) the importance of using all features along with the information content in metabolomics data, and (3) the challenge of cross-study comparison due to the host of experimental conditions and individual study goals. In light of the difficulty of cross-study comparison, we highlight limitations to the observed robustness of these results, and support efforts for standardization and data sharing.

Our analysis found multiple health states suited for biofluid-based diagnostics along with others that are challenging to diagnose. High model performance across infectious diseases suggests this category may be an attractive area for diagnostics development. This high performance may have a biological explanation, as infection involves immune response carried via the circulatory system. A similar argument could be made for renal, cardiovascular and rheumatological health states that also display high model AUCs and involve the circulatory system. Of equal importance are challenging to diagnose health states (e.g. cancer), for which the wide range of predictive performance may stem from multiple issues. Some health states—especially in their early stages—may not be reflected in biofluid metabolite levels; in the case of cancer, this may occur due to immune suppression^48^. Individual responses to a given cancer (or other disorder) may be highly variable, and even tumor region specific^49^, possibly obfuscating any chemical signal in a biofluid. Large and diverse cohorts may minimize such variability—perhaps one reason why the large, 1005-person cancer study^25^ performed so well. Finally, predicting long-term neoadjuvant chemotherapeutic response using baseline metabolomics appears challenging (AUC ∼0.5, study B1^39^) and may require additional information including electronic health records combined with other aspects of clinical machine learning^50^.

Determining which health states are unsuitable for metabolomics-based diagnostics on the basis of these results is difficult. Our goal was not to optimize individual model performance by engineering processing parameters, using optimal data set-specific transformations, or matching machine learning models to individual problems. Instead, we attempted to treat each data set as similarly as possible in order to comment more broadly on the potential of metabolomics for diagnostics. This may have led to certain data sets displaying lower than expected performance relative to reported values. For example, cancer study B5^11^ obtained high AUC values of 0.95 and 0.93, compared to our 0.82 ± 0.01 and 0.83 ± 0.01, for distinguishing colorectal cancer from non-cancerous polyps or healthy controls, respectively. Given these considerations, it is likely too early to rule out health states for which such diagnostics can be built.

Biofluid-based metabolomics provides rich diagnostic information, much of which is often overlooked—specifically, the non-significant features. As opposed to building models solely from significant features, we found that performing L1 regularization with complete feature sets yielded improved model performance. Moreover, even non-significant features were capable of providing, in select cases, robust health state discrimination. An additional benefit of the L1-LR model is its relative interpretability, as model coefficients may help identify important molecular features.

For diagnostic tests, while substantial diagnostic information is present, mass spectrometry-based metabolomics may possess a high level of information redundancy. This observation may arise due to the high dimensionality of the data generated relative to the number of blood or urine metabolites (e.g. ∼4000 in serum^51^ or ∼2700 in urine^52^). This redundancy may occur for several reasons. At the instrumental level, high-resolution mass spectrometry may possess isotope peaks that putatively hold the same information as the monoisotopic species. At the biological level, metabolites closely connected via biochemical pathways may provide information on each other. For the significant feature models, there may be substantial mutual information, as restricting models to only the top 5–100 most significant features minimally altered performance relative to using all significant features. Information redundancy may also explain why models trained via a combination of all within-study data sets did not substantially increase model performance relative to the average of models built on individual data sets (Figure 3A). One hypothesis to explain the lack of additional performance is that the features reflect a shared underlying state or are biochemically associated, despite different physicochemical properties. Thus, while these data sets come from different ionization modes, chromatography methods, or sample types, they do not appear to provide orthogonal diagnostic information.

The studies we analyzed encompassed several distinct sample types, analyzed by many chromatographic and MS techniques, making cross-study chemical or biological comparisons difficult. Given these challenges within a single health state (e.g. cancer), it was not possible to make comparisons across health states. As a result, it is unclear whether the metabolic profiles observed are health state-specific or simply general signals of illness. Thus, while it is ideal to match the proper experimental and analytical methods to the problem of interest, doing so makes cross-study comparisons more challenging and lends minimal insight for new health states for which these parameters are not known. It appears prudent to use all available routes of data collection when possible, despite the fact that different methods may generate data with overlapping information.

While GC-MS performed worse than LC-MS, this may not reflect the true capabilities of the method. Our data processing pipeline may be better suited for LC data as IPO^53^ (‘Isotopologue Parameter Optimization’) was built for extracting parameters from LC data. In addition, a larger fraction of GC-MS studies were on cancer (63%), versus 27% of LC-MS studies—and a larger number of LC studies used HR-MS, possibly supplying more information. In fact, GC-MS is more amenable to between-lab comparisons^54^ and may supply information on a different set of molecules that are critical for diagnosis.

It is challenging to ascertain the robustness of the diagnostic capabilities obtained, considering the diversity of cohorts and the possible effects of confounding variables. Many studies directly accounted for variable like age and sex in their cohort design^16,17,21,25,32–34,45^. This information was, however, not always available and to treat all studies equally, such variables were not directly modeled. While we do not expect general variables to strongly affect performance because they were typically considered during study design, their effect cannot be ruled out. More importantly, study-specific variables like medication, familial or genetic linkage, and lifestyle may also impact the results obtained. These factors were even more rarely presented and similarly not modeled. Without access to detailed individual data, it is difficult to determine what metabolomics information is truly clinically relevant from each study, particularly when all studies are treated in a similar manner. Furthermore, a number of the studies possessed small cohorts (< 30 individuals), thus the diagnostic signatures obtained may not translate to larger and more diverse populations.

Many of the presented caveats support the metabolomics community’s efforts to standardize methods across labs and studies^55,56^, and underscore the importance of open-access sharing of data. Releasing raw MS data sets—accompanied by quality control and standard samples, run order, batch numbers, and secondary MS data—could facilitate improved data correction, compound identification, and comparisons between studies. The inclusion of general and health state-specific metadata, when possible, would also be highly beneficial. This data would significantly advance the community’s ability to build off one another’s work and would expand the diagnostic capabilities of metabolomics to more challenging problems.

## Conclusion

Leveraging L1-LR models trained on all metabolite features, our analysis of biofluid metabolomics yielded diagnostic signal across nine different health states. The vast majority of studies differed in the sample type, MS instrumentation, and experimental setup. Intra-study data set combination did not significantly improve model performance, suggesting redundancy in information content across different experimental modes of analysis (column, MS ionization mode, instrument, sample type). We observed that including all features, as opposed to only statistically significant features, improved model performance. Training models in this manner not only finds relatively sparse solutions, but also leverages both statistically significant features as well as features that are not typically used. Ultimately, we find that non-significant features alone may provide high model performance in multiple health state categories. We supply all processed MS data sets, associated metadata, and code. This study advances a data-driven approach to metabolomics diagnostics that may further set the stage to improve patient care using minimally invasive tests.

## Methods

### Study data acquisition

Studies were obtained from The Metabolomics Workbench, http://www.metabolomicsworkbench.org/ or Metabolights (https://www.ebi.ac.uk/metabolights/) ^57^ with the exception of the study by Feng *et al*. that was obtained from https://datadryad.org/resource/doi:10.5061/dryad.s8k81 with additional data from their supplementary information. A full table listing the study identifiers, project IDs, and project DOIs is listed in Table S1.

### Data processing

Python 3.6.5 with scikit-learn version 0.19.1 and R 3.5.1 were used. Following data acquisition, LC- or GC-MS files were converted to .mzML or .CDF using msconvert_ee.py (if needed), a python wrapper for msconvert in ProteoWizard^58^. Select .PEG and Agilent ChemStation .D files were converted to .CDF using a wrapper (PEG_to_CDF.py or chemstation_d_to_CDF.py) of Unichrom’s ucc.exe (http://www.unichrom.com/dle.php); conversion scripts were run on a Windows operating system.

Data was then feature extracted using full_ipo_xcms.py. This converted any .CDF files to.mzData format (using cdf_to_mzData.R), selected XCMS^59^ parameters (bw, peakMin, peakMax, ppm, noise, mzdiff, binSizeObi, gapInit, gapExtend, binSizeDensity, minFrac) by averaging the results from IPO^53^ (using three independent outputs from IPO_param_picking.R, each run on a single random LC- or GC-MS file from all processable files), and finally extracted data set features (extract_features_xcms3.R with data set-specific command line flags determined via IPO; other XCMS parameters were hardcoded). Select data sets were run on XCMSOnline^60^. These included ST000063, and three of the eight data sets from ST000046. Additionally, only the negative ionization mode data for MTBLS352 (D1) was processable. For study D2, we used both the serum and the matching DBS samples together. Processing was parallelized using StarCluster (http://star.mit.edu/cluster/) on Amazon’s Elastic Compute Cloud (EC2).

Following feature extraction, output files were parsed along with additional author-provided metadata using the python jupyter notebook extracting_features.ipynb. Each study was analyzed independently; labels (0 for controls, 1 for cases) and metadata were extracted and mapped to the samples from the XCMS output. Select multiclass studies were reduced to binary problems (MTBLS315, MTBLS579, ST000381, ST000385, ST000421, ST000396 and ST000888). For ST000381, all categories other than healthy (i.e. modest, intermediate, and severe) were considered cases. For non-reducible multiclass data sets, we created data sets for each possible one-versus-one comparison. When replicates were included for a sample, only one was arbitrarily kept, minimizing bias imposed on the data.

To correct for batch effects, all data sets were transformed using the percentile normalization strategy by Gibbons *et al*^47^. If batch information could be determined, each batch was normalized separately and then combined; lacking such information, normalization was applied to the full data set (batch_correction.ipynb). Prior to normalization, missing or < 1 peak intensities were set to 1, followed by binary log transformation of the data. For select author-data sets that appeared to be log transformed, this normalization was not performed.

Within study data set combination was performed using combining_datasets_internal_to_study.ipynb. A manually curated list of combinable data sets was created and the feature matrices were concatenated, ensuring that samples for a single individual across data sets were combined into a single feature vector.

### Adduct and isotope determination

Using only the high resolution MS data sets (Table S1), we calculated the mass difference between each statistically significant feature and all others. A feature was considered an adduct or isotope if both features possessed retention times < 15 s apart, the mass difference was not zero, and was in a defined range. Significant features were assumed to be either [M-H]^-^ or [M+H]^+^ ions. All mass windows were differences relative to one of the two states:

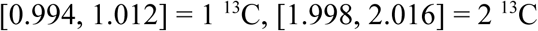

Positive ionization:

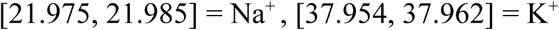

Negative ionization:

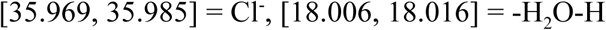

### Machine learning and statistical analysis

Model training was performed using either noncombined_model_training.ipynb or combining_datasets_internal_to_study.ipynb for the non-combined data sets and the combined data sets respectively. For each, L1-LR (sklearn.linear_model.LogisticRegressionCV, scoring=roc_auc, tol=0.0001, intercept_scaling=1 and max_iter=500) and PLS-DA models (sklearn.cross_decomposition.PLSRegression, default settings) were trained using a 5-fold outer (sklearn.model_selection.StratifiedKFold), 3-fold inner stratified and nested cross validation protocol. Inner cross validation selected either the L1-regularization parameter or the number of components used by the PLS-DA model (2, 5, 20, 50, or 100, sklearn.model_selection.GridSearchCV). Test performance was assessed on the independent fold in the outer cross-validation loop. To ensure that performance was not altered by data splits, this protocol was repeated independently 30 times on full data shuffles (sklearn.utils.shuffle) from which a data set average AUC, standard deviation and 95% confidence interval (using the average number of test cases for sample size to reflect the larger uncertainty in the smaller data sets) was calculated. The last model training was saved for analysis.

P-values were corrected for multiple testing using the Benjamini-Hochberg False Discovery Rate (BH-FDR) method applied to the results of a MW-U test (statsmodels.stats.multitest.multipletests and scipy.stats.mannwhitneyu, giving Q-values) since all multiclass problems were reduced to multiple one-to-one comparisons. To determine the number of significant features for a given data set, Q-values were calculated using the full data matrix and all values < 0.05 were considered significant. However, to train models using only statistically significant features, but not have information from the test set leak into the model training, we calculated Q-values internal to the model training loop after the 5-fold stratified data splitting; significant features found in the training step were subsequently used for testing and often differed across folds and data set shuffles. To train models using up to the *x* most significant features, a similar protocol was followed and only the *x* most significant features (largest negative log Q-values) were retained for model training and testing. If fewer than *x* features were found significant, only those features were used, despite not reaching the specific value of *x*. For models trained with only non-significant features (Q-values ≥ 0.05), all significant features from the complete full data set were removed prior to training.Features belonging to adducts and isotopes were similarly removed prior to training; such models were only built for the non-combined, high resolution MS studies. If any data set possessed 0 features, an AUC of 0.5 was recorded and training stopped. Overfit models with test AUC values less than 0.5 were retained. For each data set, feature coefficients from trained models were averaged.

MW-U tests were used for the significant testing of: GC and LC, positive and negative ionization, C18 and HILIC, plasma and serum, as well as all one-to-one ‘all features versus *x* significant features’ comparisons. Feature enrichment was calculated for each percentile-normalized feature as the mean value in the cases divided by the mean value in the controls.

## Data Availability

All mass spectrometry data used is freely available and listed in Table S1. Packaged data sets .pkl and input feature with matched label .csv files, along with model training output .csv and .pkl files will be deposited in Zenodo upon publication.

## Additional Note

^‡^Refers to data sets without a published accompanying manuscript. This includes the following studies: ST000329, ST000763, ST00062/3.

## Code Availability

All code (.sh, .py, .R, and .ipynb) will be uploaded to the following Github repositories upon publication: https://github.com/ethanev/Metabolomics_ML and /Metabolite_lookup.

## Acknowledgements

We would like to J. Zhang for his helpful comments and insights. This work was supported by The Abdul Latif Jameel Clinic for Machine Learning in Health at MIT (J-Clinic) as well as the Center for Microbiome Informatics and Therapeutics (CMIT).

## Author contributions

EDE processed the data and performed the analysis. CD, NDC and MKO provided valuable discussions and suggestions. MAM and IR helped create the study list and collect data. EJA and DS provided insight on the project direction and manuscript. EDE and EJA wrote the manuscript and all authors edited or approved of the manuscript.

